# Overcoming COVID-19 vaccine preferential bias in Europe: Is the end of the pandemic still foreseeable?

**DOI:** 10.1101/2021.06.23.21259422

**Authors:** Frank Adusei-Mensah, Ivy Eyiah Inkum

## Abstract

The availability of safe and effective vaccine alone does not save lives; it is the inoculation plus other public health measures that do. Recent reports suggest the growing trend in vaccine preferential bias in parts of the world but not much in Europe. The present paper aims to investigate the occurrence of COVID-19 vaccine preferential bias in Europe for effective vaccination planning and pandemic control.

**Method:** Data on vaccine delivered and vaccination campaigns of the EU member states was collected from Eu center for disease control (EUCDC) on COVID-19 vaccination radar. The data was processed for analysis on MS excel and both descriptive and statistical analysis was done with IBM’s SPSS version 21. Analysis was performed at 95% confidence interval and statistically significant difference was considered at p < 0.05.

**Results:** We observed statistically significantly lower vaccine uptake compared to the vaccine delivered doses in the present study (average at 62.678 +/-3.928%) (p< 0.05, CI = 95%). Great variances in uptake for Oxford-AstraZeneca vaccines (50.927 +/-4.626 %) compared to Pfizer-Biontech vaccine (86.285 +/- 2.1052 %) was observed compared to previous prospective study on the wiliness to receive COVID-19 vaccine in the region (75%).

**Conclusion:** Public health practitioners and policy makers need to factor the existence of COVID-19 preferential bias based on vaccine type or manufacturer. This will enable them introduce policies including public educational campaigns to overcome biasness on the wiliness to inoculate thereby enhancing vaccine uptake for smooth and effective control of the pandemic.

## introduction

Coronavirus pandemic is a global challenge affecting the lives and livelihood of billions globally. The unprecedented speed of the pandemic tested every ounce of science, and it has led to the development of vaccines by different vaccine makers at an unprecedented pace than the world has ever experienced before. Being one of the most important advances in modern medicine, vaccines have saved the lives of many from vaccine preventable deaths. Credit to the success of vaccination, some of the world’s devastating pandemics including smallpox has been controlled and eradicated. Vaccination is undoubtedly one of the efficacious public health arsenals of modern medicine. However, it is important to note that having safe and efficacious vaccines alone do not save lives from pandemic; but rather, vaccination plus other public health measures do! The scale of the pandemic requires Widespread vaccination as an indispensable element for controlling the COVID-19 transmission. Although there still remains unanswered questions about the degree of protection, the duration of the warning of the vaccine triggered immunity to COVID-19 begins [1]. With over 175.3 million vaccine doses administered between Early December 2020 to 15^th^ February 2021 globally, there is much to be desired for effective control and eradication for the COVID-19 pandemic [2]. To achieve efficient vaccination campaign, public health professionals and vaccination planners need to overcome many bottlenecks including perceptions on vaccine safety, side effects and efficacy [3]. However, amidst a backdrop of extensive mistrust in the effectiveness and safety of coronavirus vaccines globally, the COVID-19 is still spreading at an unparalleled speed [1]. With vaccine producers stretched, production and supply chains tested, relying on a single producer alone will prolong the pandemic. Since most of these vaccines available have impeccable results in averting severe covid-19 infection and mortality, it is advisable to say that the best vaccine to safe one’s life is the one that reaches him/her in time. Most recently, there have been reports of COVID-19 vaccine bias with many preferring one vaccine to the other. With thousands across Europe cancelling vaccination appointment times preferring Pfizer to AstraZeneca/Oxford University vaccines and close to 80% of the Oxford/AstraZeneca vaccine doses delivered to EU countries remain used for reasons of efficacy, safety and region of manufacturing [4]. With billions of lives at risk and tens of thousands of lives lost every day to COVID-19, it is important to evaluate the vaccines available today for their efficacy and safety from epidemiologic point of view and to promote vaccine uptake. The main aim of this paper is therefore to collate the available information on the vaccines and to draw the public attention to the need to reduce being vaccine bias for effective control of the pandemic.

### Attitude to get vaccinated

As it is known that vaccines do not save lives; vaccination does. Nearly 2 decades after the approval of rotavirus vaccine by the FDA and the WHO, yet only about 60% of the world’s children have received complete dose of the rotavirus vaccine [5]. COVID-19 vaccine preferences, hesitancy and an ‘access gap’ would have dramatic consequences on lives [5]. There are questions that are common to vaccines globally; access issues, logistics and supply chain management, and vaccine administration [5]. The scale and urgency of the current pandemic makes it even more dicey, requiring continues research to promote acceptance rate. Briefly, some of these several factors that may thwart the use of COVID-19 vaccines include: The programmatic scale for different working and age groups, logistics with consideration on cold-chain facilities for mRNA vaccines of Moderna (−20□°C, lasting maximum 7-days) and Pfizer (−70□°C lasting maximum 3 days) which are exclusively thought-provoking issue for use in Low- and Middle-income countries (LMICs). Again, of particularly important in Europe is the acceptability of the various COVID-19 vaccine platforms under mass use. In a previous study, acceptance in the region was about 75% [6]. But this was a prospective study without the existence of various platforms with varying phase III efficacy and safety profiles.

Of great interest of vaccine acceptability is the public’s attitude towards vaccine safety, their importance, and effectiveness. Distrust in the safety and effectiveness of vaccines have been consistently associated with vaccine uptake [7]. Although the general population of Europe have previously indicated positive attitudes towards vaccines [1], they did not indicate which vaccines types or platforms they would like to be vaccinated with. Previous study suggests close to 26% of the adults population across seven European countries were uncertainty and unwilling to get a COVID-19 vaccine when available [8], the real figures in practice are much higher favoring certain vaccine platforms [4]. In the same study, about 70-80% of Europeans were ready to receive the COVID-19 vaccine when available [8]. Specifically, 73.9% of the participants (n=7664) from Denmark, France, Germany, Italy, Portugal, the Netherlands, and the UK were willing to get vaccinated against COVID-19. Though, Neumann-Böhme and colleagues identified fear of possible side effects of the future COVID-19 vaccines as a major reason for not wanting to be vaccinated against COVID-19, it has become clear that most of the riot in Europe towards COVID-19 vaccine preferences are related to efficacy. The above poses significant challenge in achieving the high vaccination coverage required to control the pandemic and return to near normal life.

With the global supply and production chain stretched to the limit and scarcity of raw materials to meet high vaccine demand, it is important to evaluate the available vaccines to see if it is worth being choosy and picky or accepting whichever vaccine made available to you to help control the pandemic.

### Characteristics of the COVID-19 vaccines in mass administration in Europe

The Pfizer/BioNTech and Modena are mRNA vaccines expressing the COVID-19 spike glycoprotein. In principle, these vaccines use the body’s DNA to translate the genetic message from the COVID-19’s viral mRNA into the spike glycoprotein’s De novo synthesis. The translated spike glycoprotein then induces the body’s immune response against future viral infection. However, the vaccines from Gamaleya and AstraZeneca-Oxford University express the spike protein from adenovirus vector platforms. In principle, the AstraZeneca vaccine uses a spike protein expressed from chimpanzee’s adenovirus [5]. It is worth noting that, effective vaccination is a part of active, all-inclusive control of a pandemic [5]. Data from the AstraZeneca’s vaccine phase III trial suggest that, the overall vaccine efficacy 14 days after the second dose was 66·7% (ChAdOx1 nCoV-19), **(Table 1**), and no hospital admissions was observed after the initial 21-day [9]. Less than 1% of the 12,282 participants had serious adverse events but no vaccine related death observed [9]. They had a tentatively efficacy of 76·0% with a single standard dose between 22 to day 90 days after vaccination [9]. The Pfizer BionTech vaccine reported 94% - 95% efficacy in their phase three trials after their second dose administered weeks apart whiles Modena reported similar efficacies with similar dose variations [10]. The Modena’s mRNA-1273 vaccine against COVID-19 encrypting for a prefusion stabilized form of the Spike (S) protein[11]. According to the WHO report, the Modena vaccine has efficacy of approximately 92% in protecting against COVID-19, 14 days after the first dose [12], **(Table 1)**.

**Table 1:**
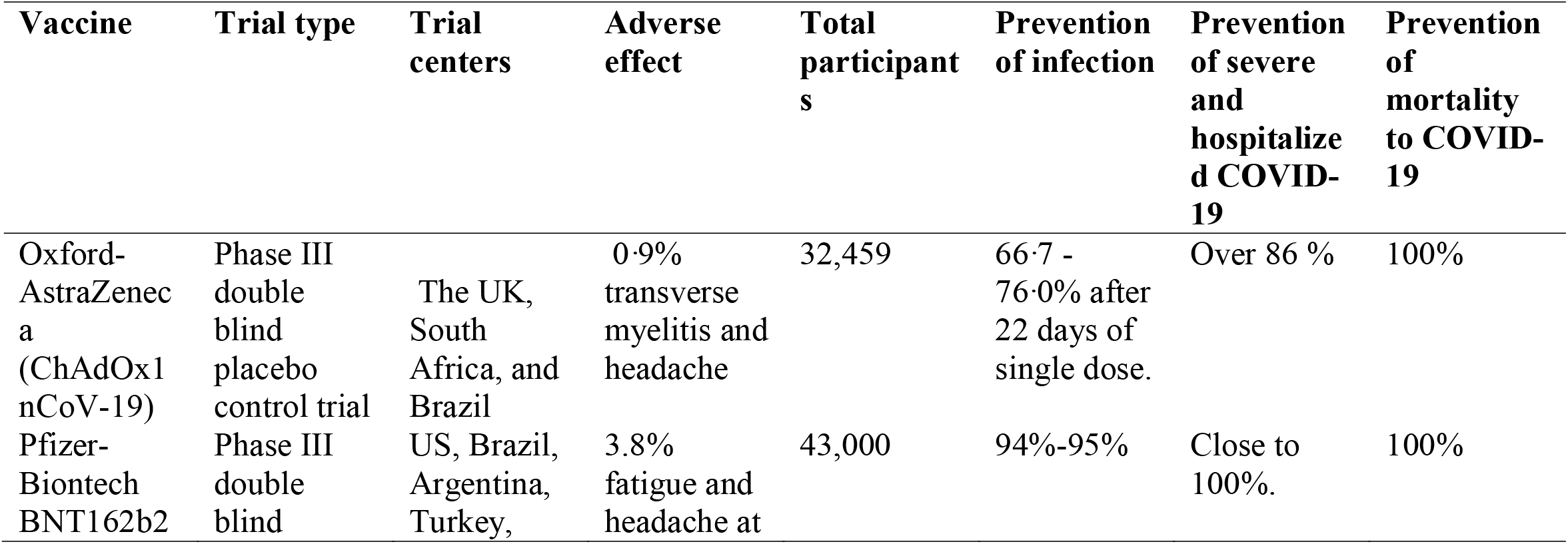
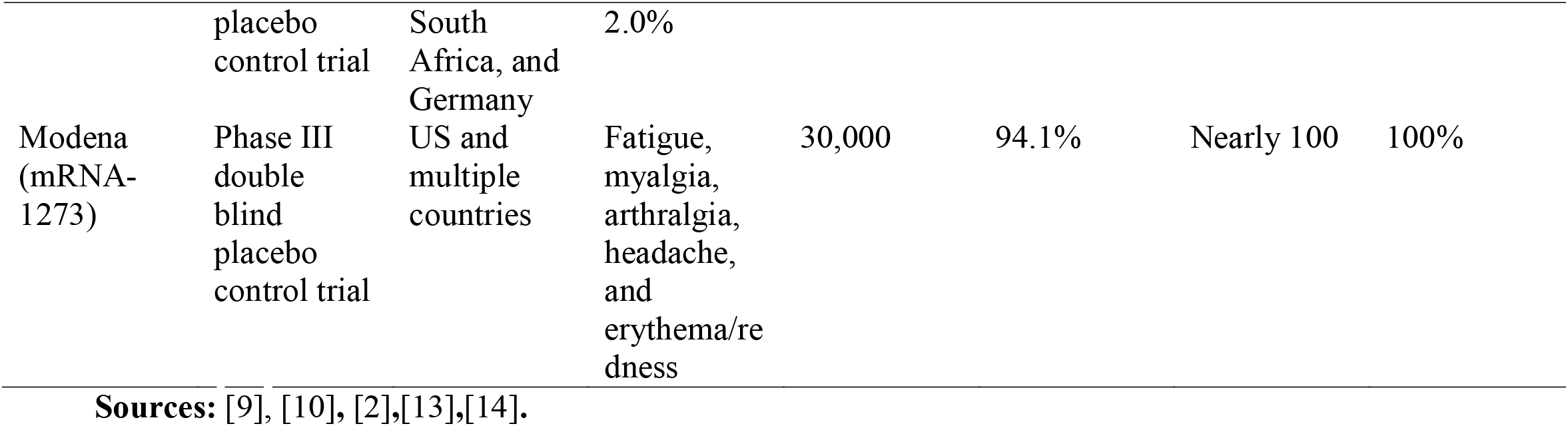
Basic information on the COVID-19 vaccines in mass use.

## Methods

Vaccination data in the European region on the EU member states was obtained from Eu center for disease control’s vaccine tracker (EUCDC) [15]. The data is publicly available for academic and non-for-profit use. However, the authorities were informed through email communication on the intent to use the data. As a result, all ethical or legal requirements for the use of the data was met. The extracted data was organized for analysis using Microsoft’s Excel 2010. Descriptive and statistical analysis were performed using IBM’s SPSS version 21 and analysis at 95% confidence interval and statistically significant difference was considered at p < 0.05.

## Results

Data was obtained from European Centre for Disease Prevention and Control, COVID-19 vaccine tracker **[15]**. The difference between the total vaccines distributed to the member states (n=**47**,**319**,**245)** and the vaccines administered by the members (n=**34**,**827**,**459) as of** by 5^th^ March 2021 was significantly low (p < 0.05) at 95%CI (p=0.00) **[15]**. Though there have been production challenges by vaccine producers in meeting the huge demand, the figure 1 presents a clear difference in the doses received compared to the doses administered in the EU region. The received-administered differences vary between countries but very conspicuous in countries like France, Germany for AstraZeneca vaccines for AstraZeneca vaccines (Figure 1). The overall administered by platform was observed to be low as 50.927 +/- 4.6258 SE for AstraZeneca, 50.822 +/- 5.0502 SE for Modena and as high as 86.285 +/- 2.1052 SE for Pfizer/Biontech vaccines (Table 2 figure 2).

**Figure 1:**
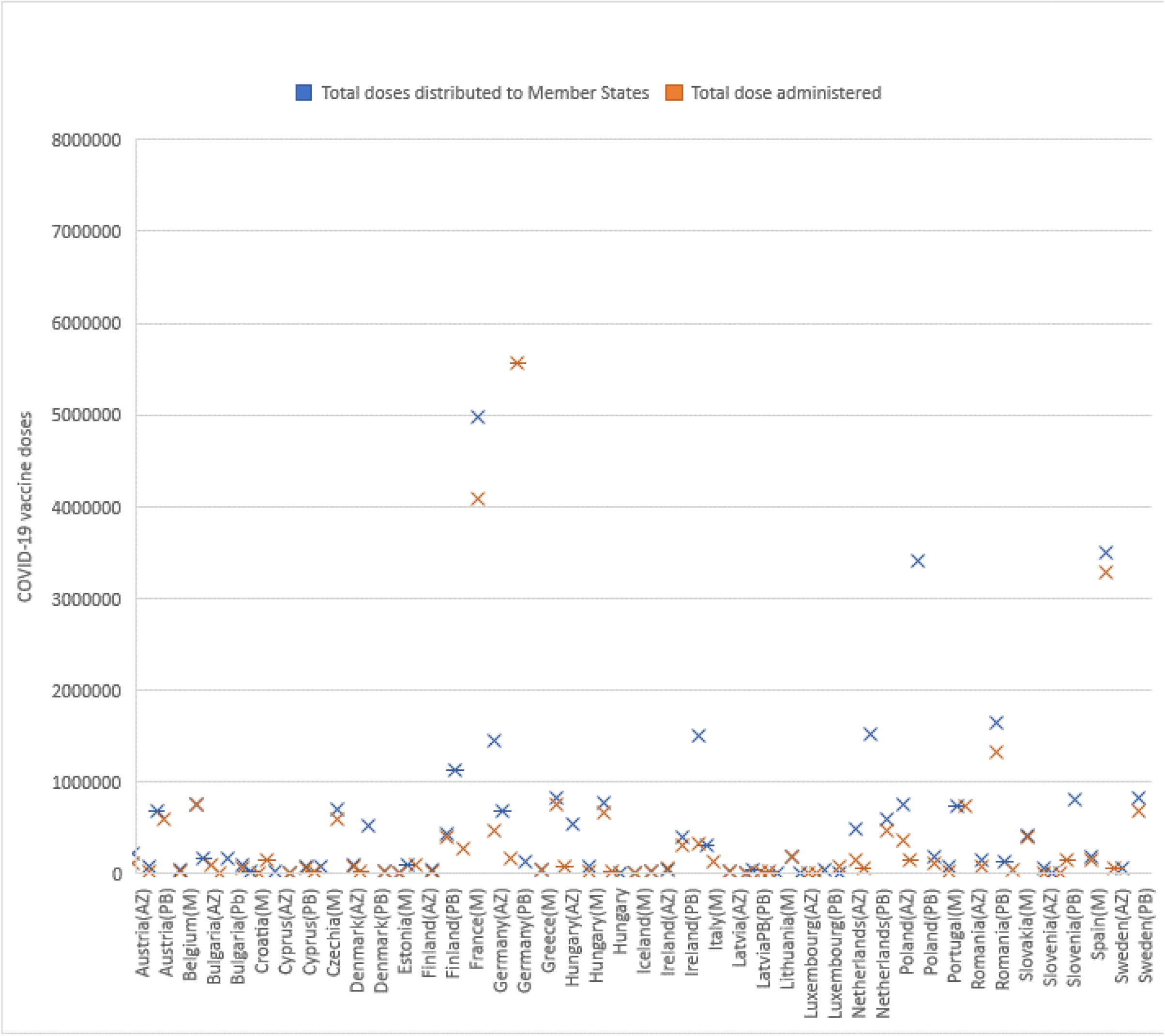
COVID-19 vaccine doses received and administer in Europe as at 05^th^ March 2021. Note: AZ = AstraZeneca, PB = Pfizer-Biotech and M = Modena.

**Table 2:** ANOVA analysis compared to the vaccine doses received.

|  |  | Sum of<br>Squares | df | Mean Square | F | Sig. |
| --- | --- | --- | --- | --- | --- | --- |
| Total dose<br>administered | Between<br>Groups | 77145352756<br>208.100 | 82 | 94079698483<br>1.806 | 20902.012 | .000 |
|  | Within Groups | 90019754.500 | 2 | 45009877.250 |  |  |
|  | Total | 77145442775<br>962.600 | 84 |  |  |  |
| Vaccine Categories | Between<br>Groups | 149.294 | 82 | 1.821 | .910 | .662 |
|  | Within Groups | 4.000 | 2 | 2.000 |  |  |
|  | Total | 153.294 | 84 |  |  |  |

| Product | Mean Statistic<br>+/- (Std. Error of<br>mean) | 95%<br>Confidence<br>Interval |
| --- | --- | --- |
| AstraZeneca | 50.927 +/- 4.6258 | 41.419 - 60.436 |
| Moderna | 50.822 +/- 5.0502 | 40.441 - 61.202 |
| Pfizer/Biontech | 86.285 +/- 2.1052 | 81.957 - 90.612 |
| Average % | 62.231 +/- 2.9438 |  |

**figure 2:**
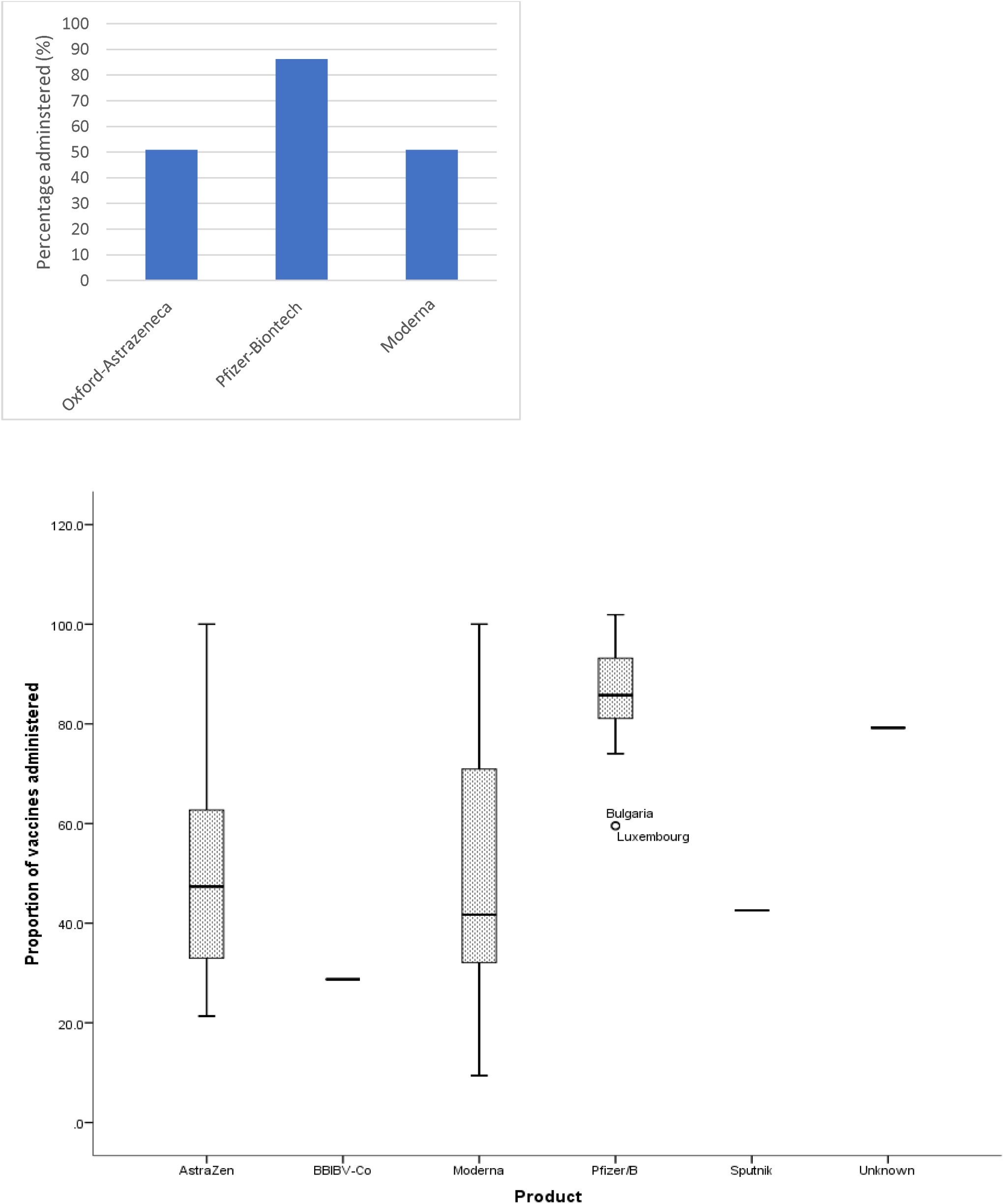
Descriptive statistics of the percentage of vaccines administered

**Figure 3:**
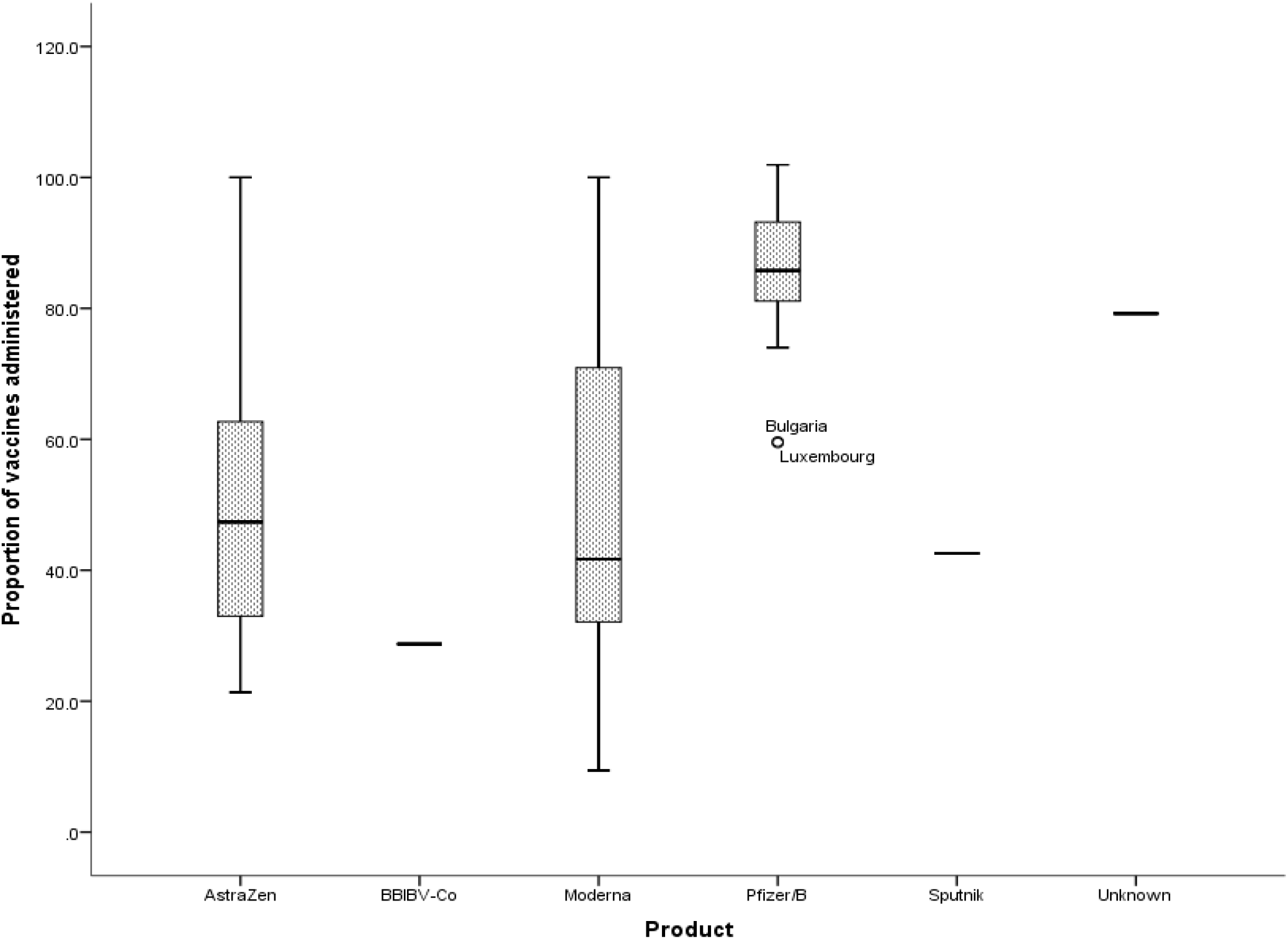
Stem plot. The stem plot shows the mean statistics and the deviations of the various vaccine platforms in the region. There exists great variation in the administered Modena and AstraZeneca vaccines in the region compared to that of the Pfizer-Biontech vaccines (Figure 3).

**Figure 4:**
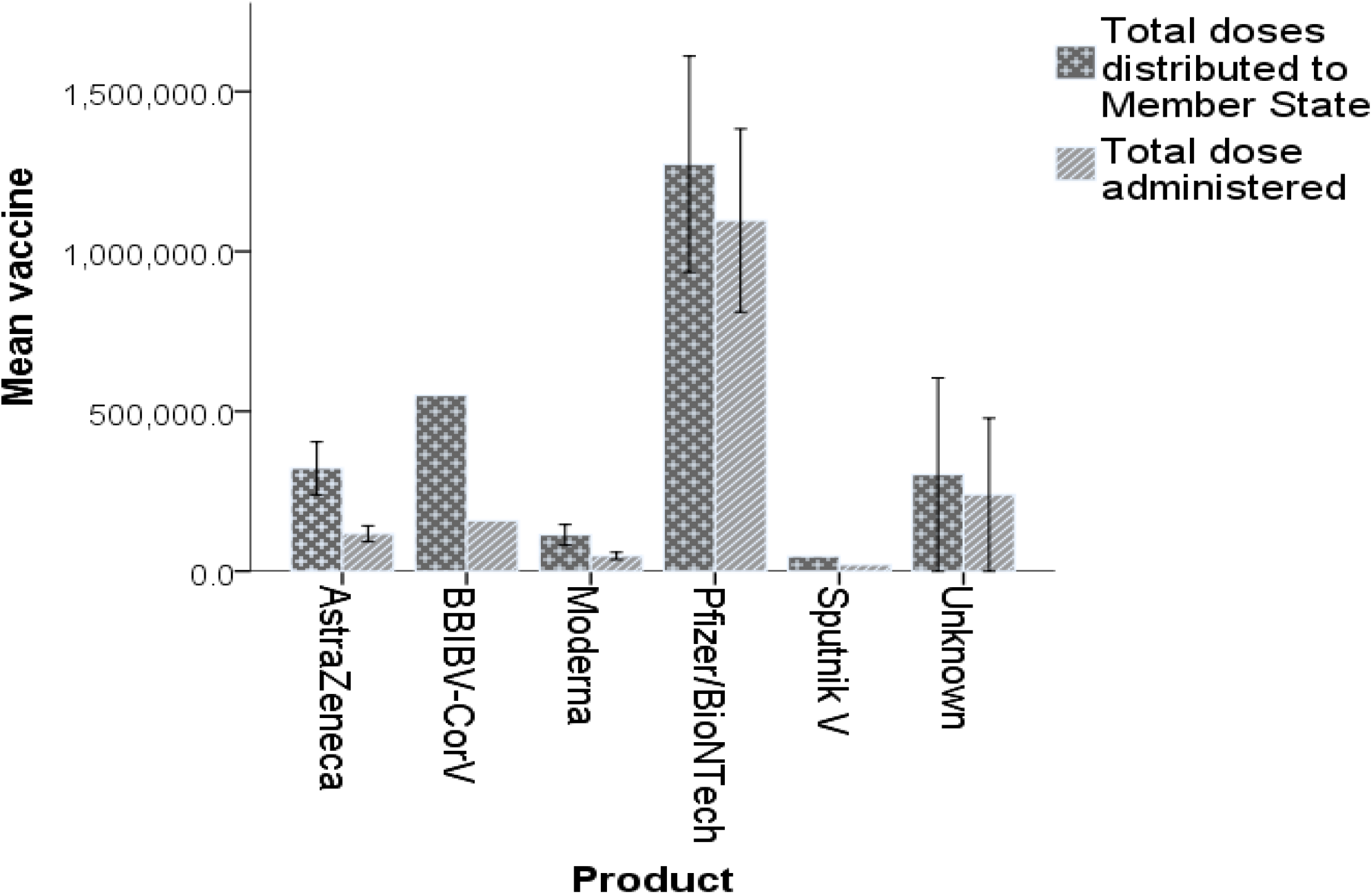
Administered vaccines. Over 1 million Pfizer vaccines have already been administered in the region while the acceptance of AstraZeneca, BBIBV-CorV vaccines administered compared to doses distributed are on the low side (Figure 4).

There exists a significant difference between the received and the administered vaccines by the member states. Compared to distributed doses, the administered doses were significantly lower than the control (p=0.00) at 95% CI table 3.

The acceptance of the COVID-19 vaccines are very high in countries like Denmark (100%) and Lithuania (100%) with 100 percent utilization of all the received vaccines. On the centrally, the administered rates are low in countries including Poland, France, Germany, Italy, and Spain. These four countries accounts for over 50% of the unused vaccines in the Eu member states (Figure 5). This findings confirms the previous report on COVID vaccine hesitancies[4], [6],[16].

**Figure 5:**
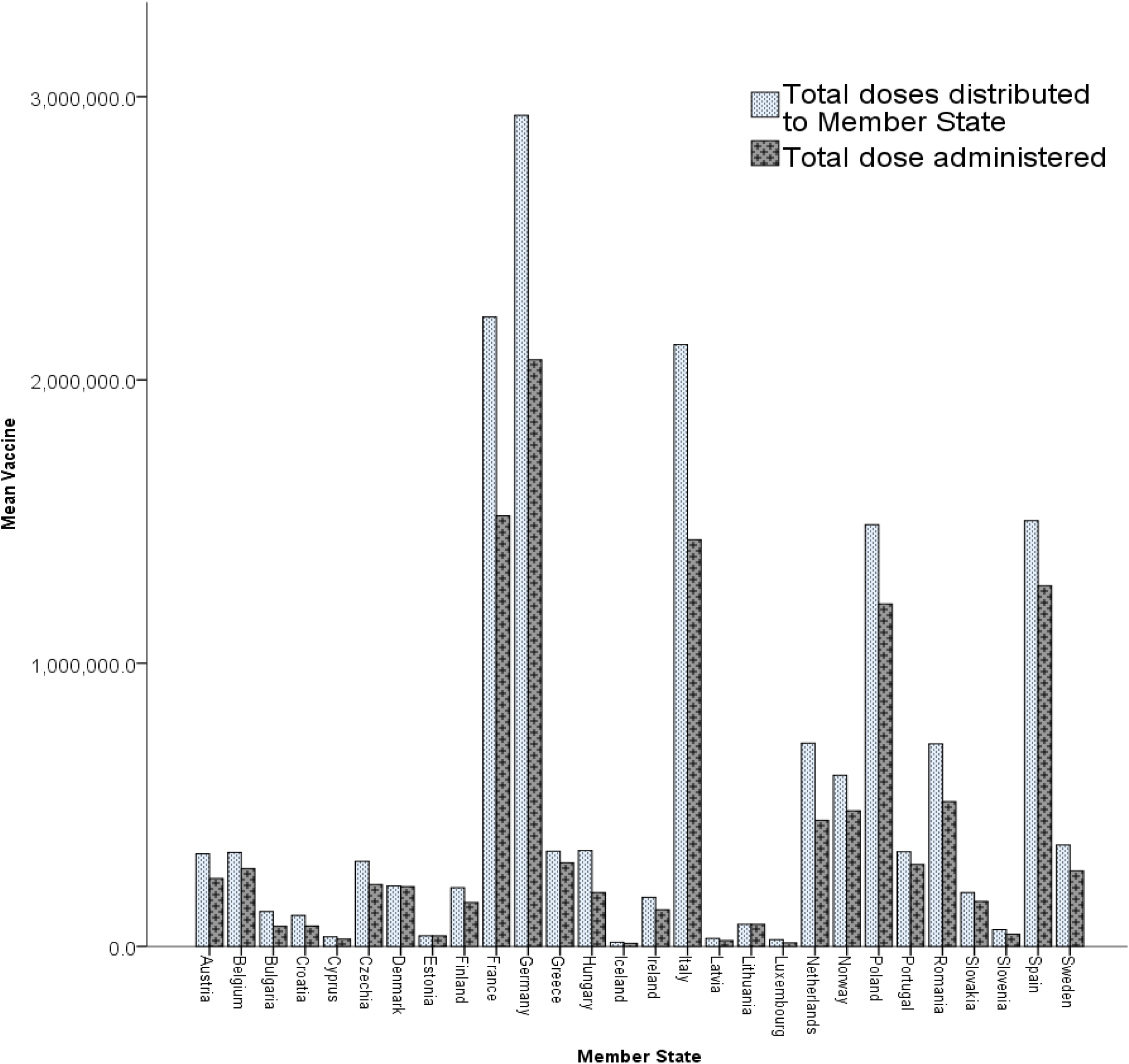
Country performance of the distributed and administered vaccines.

## Discussion

Defined by the WHO, ‘Vaccination is a simple, safe, and effective way of protecting people against harmful diseases, before they come into contact with them’’. [17]. Vaccines uses the body’s natural immune system to build resistance to specific infections prior to contact to the pathogen. As important as vaccines are, it must be made clear that having safe and effective vaccines alone to not guarantee protection, it is the immunization plus other health measures that ensures protection. It is therefore important therefore to attach of particular importance to factors that influences vaccine uptake. There has been numerous studies on vaccine hesitancy and acceptability in the past and factors such as trust of the vaccine’s safety and efficacy are of particular importance [6], and these factors are shared globally [18][19]. These previously identified factors need a more critical consideration by policy makers to promote vaccine uptake. It must be stated that since the vaccine trials were not initially planned head-to-head, a mere comparison of the clinical trials alone of the various vaccine platforms would be a great mistake. These trials were performed at different phases of the pandemic either before or during the confirmation of the variant strains such as the B.1.1.7 of the UK and South Africa, the B.1.351 of the Brazil, California which are more virulent than the formal [20]. It is worth noting that all these vaccines are doing best what they ought to do; that is to be safe and to activate the immunes system to fight against diseases and death. People have experienced flu after receiving flu shots in the past, however the assurance after receiving flu vaccine is protection from serious flu during the flu season. Similarly, the current corona vaccines that are safe and nearly 100% assurance of protection from serious COVID-19 and or mortality from the disease should receive similar attention if we would like to return to the near normal in the shortest possible time.

Findings from the present study shows that the acceptance of the COVID-19 vaccine is generally lower than predicted in previous studies that investigated the wiliness of acceptance COD-19 vaccines if they are available. Average acceptance rate of 62.231% +/- 2.9438% was observed in the present study which is lower than reported previous studies. In a large global survey published in nature, 71.5% of participants (n=13,426) reported their wiliness to take a COVID-19 vaccine when available while 48.1% reported of taken it based on their employer’s recommendation [21]. They however observed differences in acceptance rates ranged from almost 90% (in China) to less than 55% (in Russia). High acceptance rate of about 90% in the China and Indonesia has been reported in different than France, 78% [22][23] and other parts of the world [21][23]. In a multi-state European survey, acceptance rate of 73.9% (7664) has been reported with participants from Denmark, France, Germany, Italy, Portugal, the Netherlands, and the UK [24]. The observed lower acceptance rate of 62.231% +/- 2.9438% in the present study (Table 2, Figure 1) is lower compared to the reported wiliness rate (73.9%) to vaccinate with COVID-19 vaccines prior to the availability of the vaccines in the region.

High differences in the acceptance rate were also observed in the present study between the vaccine variants. Among the most popular vaccines in the region during the study, a high acceptance rate of 86.285 +/- 2.1052% was observed for Pfizer/Biontech vaccine while lower rates of 50.927 +/- 4.6258% for AstraZeneca vaccine and 50.822 +/- 5.0502% for Modena and less than 50% for BBIBV-CorV vaccine (Table 2) were observed. It could be observed that Pfizer/Biontech vaccine received higher uptake than the pre general COVID-19 vaccination surveys in the region 77.3% [24]. However, the rates observed in the present study for both Modena and AstraZeneca is much lower (about 51%) compared to previous pre-vaccination surveys [24][25]. Major factors such as misinterpretation and unfair-comparison of the clinical trials data of the present major vaccines in the region, conspiracy beliefs that foster mistrust, efficacy and fear of both long- and short-term side effects could be responsible for the lower vaccine up-take observed in the present study. The biasness in the uptake observed in the present study between Pfizer/Biontech and the other vaccine groups could be hugely due to the differences in their phase’s III safety and efficacy data. It must be noted that, both Modena and Pfizer/Biontech are RNA vaccines, but the uptake is higher for Pfizer/Biontech which recorded phase III trial efficacy of about 95% compared to 94% for Modena (Table 1). Again, Pfizer/Biontech also recorded a 100% in averting server COVID-19 in its 43,000 participants in the phase III trials.

Country comparison in the present study (Figure 5) shows lower utilization rates for countries like Poland, France, Germany, Italy, and Spain accounting for nearly 50% of all unused vaccines. Previously, lower rates of 53.7% for Italy [26], 58.9% for France [21] and 68.4 for Germany [21] have been reported which corresponds to the rates observed in the present study.

It is worth noting that, the clinical trials of some COVID-19 vaccine candidates were put on partial hold due to the report of a serious adverse event [27]. While there are concerns of the lack of or insufficient publicity of trial data or extremely limited data of some candidates including the Russian Sputnik vaccine prior to general public use [28]. Again, an area of consideration is the need to clear the misconception of the growing concern about future risk of worsening diseases’ pathogenesis through antibody-dependent enhancement (ADE) after immunization which was observed in the SARS-COV 1 epidemic’s vaccine candidates in *in vitro* studies [29][30]. However, the risk of ADE is usually picked up during pre-clinical and large scale clinical trials phase observed as could be observed in HIV-1 vaccine candidate [31], FIV vaccine candidates [32] and EIAV vaccine candidates [33]. It must be stated that, proper adjuvant selection may significantly influence the vaccine’s associated ADE risks.

## Conclusion

Vaccines train the immune system to create antibodies a particular disease. Though data on how long the obtained immunity from the various COVID-19 vaccine platforms will wan/persist are lacking, the available data indicate their ability to safely protect the population from serious COD-19 and mortality from the disease. Misinformation and misunderstanding of the incompatibility of the various phase III trials has led to rejection and low acceptance of certain COVID-19 vaccine platforms in the EU region. As a fact, vaccine hesitancy remains an insistent global threat. The present study observed negative preferential bias towards AstraZeneca and Modena vaccines (50% acceptance rate) compared to Pfizer vaccines (over 80% acceptance rate) in the EU region. The finding shows there exists a significant challenge in achieving the high vaccination coverage required to control the pandemic and return to near normal life. These call for structured health education in order to promote vaccine uptake and to promote effective control of the pandemic is urgent. These observations are of concern and needs much consideration since it slows down the control measures and the lingering of the pandemic. Such educations should put some level of emphasis on the safety, efficacy, sufficiency, and availability of Trials data and education on the ADE as reported in various viral vaccine trials.

## Data Availability

All necessary data have been included

